# Distinct early IgA profile may determine severity of COVID-19 symptoms: an immunological case series

**DOI:** 10.1101/2020.04.14.20059733

**Authors:** Christine Dahlke, Jasmin Heidepriem, Robin Kobbe, René Santer, Till Koch, Anahita Fathi, My L. Ly, Stefan Schmiedel, Peter H. Seeberger, ID-UKE COVID-19 study group, Marylyn M. Addo, Felix F. Loeffler

## Abstract

SARS-CoV-2 is the causative agent of COVID-19 and is a severe threat to global health. Patients infected with SARS-CoV-2 show a wide range of symptoms and disease severity, while limited data is available on its immunogenicity.

Here, the kinetics of the development of SARS-CoV-2-specific antibody responses in relation to clinical features and dynamics of specific B-cell populations are reported. Immunophenotyping of B cells was performed by flow cytometry with longitudinally collected PBMCs. In parallel, serum samples were analyzed for the presence of SARS-CoV-2-specific IgA, IgG, and IgM antibodies using whole proteome peptide microarrays. Soon after disease onset in a mild case, we observed an increased frequency of plasmablasts concomitantly with a strong SARS-CoV-2-specific IgA response. In contrast, a case with more severe progression showed a delayed, but eventually very strong and broad SARS-CoV-2-specific IgA response.

This case study shows that determining SARS-CoV-2-specific antibody epitopes can be valuable to monitor the specificity and magnitude of the early B-cell response, which could guide the development of vaccine candidates. Follow-up studies are required to evaluate whether the kinetics and strength of the SARS-CoV-2-specific IgA response could be potential prognostic markers of viral control.

## Background

The novel coronavirus SARS-CoV-2 was first described in Wuhan, China, in January 2020, as the causative agent of COVID-19.^1^ Three coronaviruses, including SARS-CoV-2, are known to have crossed species barriers to infect humans.^2-3^ Besides SARS-CoV-2, SARS-CoV and MERS-CoV are pathogenic in humans, can cause severe illness and death, and have the potential for epidemic and pandemic spread. The World Health Organization (WHO) has classified these three viruses as priority pathogens to accelerate the development of vaccines and therapeutics to prevent epidemics.

SARS-CoV-2 is a beta-coronavirus and shows genetic similarity with SARS-CoV. SARS-CoV-2 is highly infectious and is spreading fast, currently at pandemic proportions. In the four months since initial detection, more than 1.5 million people have been infected globally, while more than 89 900 have died.^4^ The infection presents with different symptoms and a wide spectrum of severity.^5^ Some patients do not have fever, cough, or radiologic abnormalities on initial presentation. Some patients only experience fever and cough, while others display the very severe form of the disease that leads to bilateral pneumonia, which has to be treated in intensive care units, and sometimes leads to death.

A better understanding of the immunogenicity and pathobiology of SARS-CoV-2 infections in humans is urgently needed as a basis for the development of diagnostics, therapeutics, and vaccines against SARS-CoV-2.

High-density peptide arrays enable the rapid identification of antigen epitopes, recognized by antibodies for many applications.^6^ Pathogen-specific peptide arrays help to identify biomarkers for (early) detection of diseases,^7^ to develop therapeutics, or to rationally design vaccines. Identification of epitopes that are targeted by protective antibodies is critical to develop monoclonal antibodies as therapeutics.

Using high-density peptide arrays in combination with clinical and immunological data provides insight into immunogenicity induced by SARS-CoV-2 with a focus on antibody development during infection. Screening patient-specific IgA, IgG, and IgM responses to the full viral proteome, mapped as overlapping linear peptides on an array, is fast and provides data to identify new diagnostic markers, and to shed light on unknown immunological interactions.

Here, we report longitudinal B-cell population and virus-specific antibody response data from two SARS-CoV-2-positive patients: a married couple. While the husband had a more severe course of disease and was hospitalized (no ventilation), his wife began to experience only mild symptoms six days after her husband and was subsequently diagnosed. In addition to identifying biomarkers for disease progression, we discovered important epitopes for vaccine development.

## Methods

### Patient Material

Blood from four patients and one healthy control were collected at the University Medical Center Hamburg-Eppendorf. Donors provided written informed consent. The protocol was approved by the Ethics Committee of the Hamburg Medical Association, Germany. Peripheral blood mononuclear cells (PBMCs) were isolated and cryopreserved from EDTA-blood using standard operating procedures via Ficoll density gradient centrifugation. Serum samples were aliquoted and stored at −80°C (see Table 1).

**Table 1.**
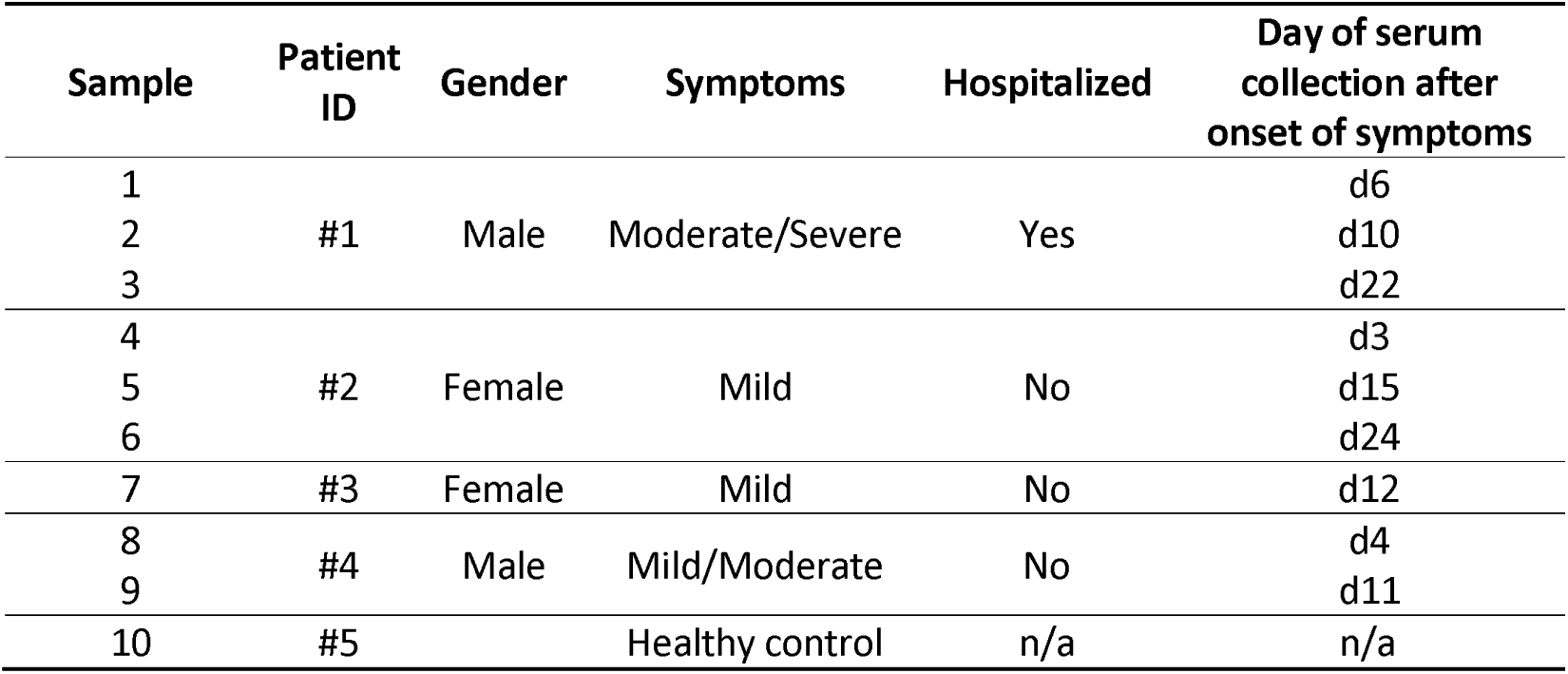
Patient and serum sample information.

### Immune-phenotyping

B-cell immune-phenotyping was conducted on cryopreserved PBMCs. Staining panels are depicted in the Supporting Information (Table S1). Cells were analyzed on a BD LSRFortessa and evaluated with FlowJo10.

### Peptide microarrays

The whole proteome of SARS-CoV-2 (GenBank ID: MN908947.3) was mapped as 4 883 spots of overlapping 15-mer peptides with a lateral shift of two AA on peptide microarrays, obtained from PEPperPRINT GmbH (Heidelberg, Germany). Patient sera were diluted 1:200 and incubated on the arrays overnight. Afterwards, IgG, IgM, and IgA serum antibody interactions were differentially detected with fluorescently labeled secondary antibodies. For details, see Supporting Information.

## Results

We collected blood at different time points post symptom onset (Table 1) and used flow cytometry and peptide microarrays to evaluate in detail, the kinetics of antibody development.

Patient #1, a 64-year-old male, developed general weakness, myalgia and headache, intermittent episodes of very high fever and subsequently, a productive cough. Two days after the first symptoms, he tested positive for SARS-CoV-2 by RT-PCR. At that point in time, fever had already subsided, but a low grade temperature recurred in the second week. Because of an increase of C-reactive protein (53 mg/dl), oral treatment with a beta-lactamase antibiotic was started. The patient was hospitalized for four days, and showed moderate but typical ground glass opacities on high-resolution thorax computed tomography scan; he fully recovered without ventilation support. Even though the patient did not require intensive care treatment or ventilation and the symptoms were moderate/severe, we define this patient as a (more) severe case than our other patients, due to hospitalization. Patient #2, his wife, a 62-year-old female, was tested SARS-CoV-2 positive six days after her husband’s first symptoms. She had high viral shedding of SARS-CoV-2 monitored by RT-PCR, although she reported only very mild clinical symptoms of COVID-19, such as sub- febrile temperatures, a mild cough, and a constant sense of well-being (Pfefferle et al.; submitted). Patients #3 – #5 were included as controls: While #3 and #4 tested positive for SARS-CoV-2, with mild (#3) and mild/moderate (#4) courses of disease, without hospitalization, sample #5 served as negative control.

The dynamics of the immune response were analyzed by flow cytometry using PBMCs. The memory B-cell population (CD19+CD24+CD38-/low) increased, following infection after approximately 15 days post disease onset in both patients #1 and #2 (Fig. 1a, b), and persisted in the severe case (#1) until d32. An expansion of plasmablasts (CD19+CD27+CD38+) in the blood circulation was detected in the mild case very early at day 3 post onset of symptoms (Fig.1b), before symptoms subsided. The severe case (#1) showed an increase in plasmablasts as the symptoms began to resolve. Early time points were not analyzed by flow cytometry from this patient.

**Figure 1.**
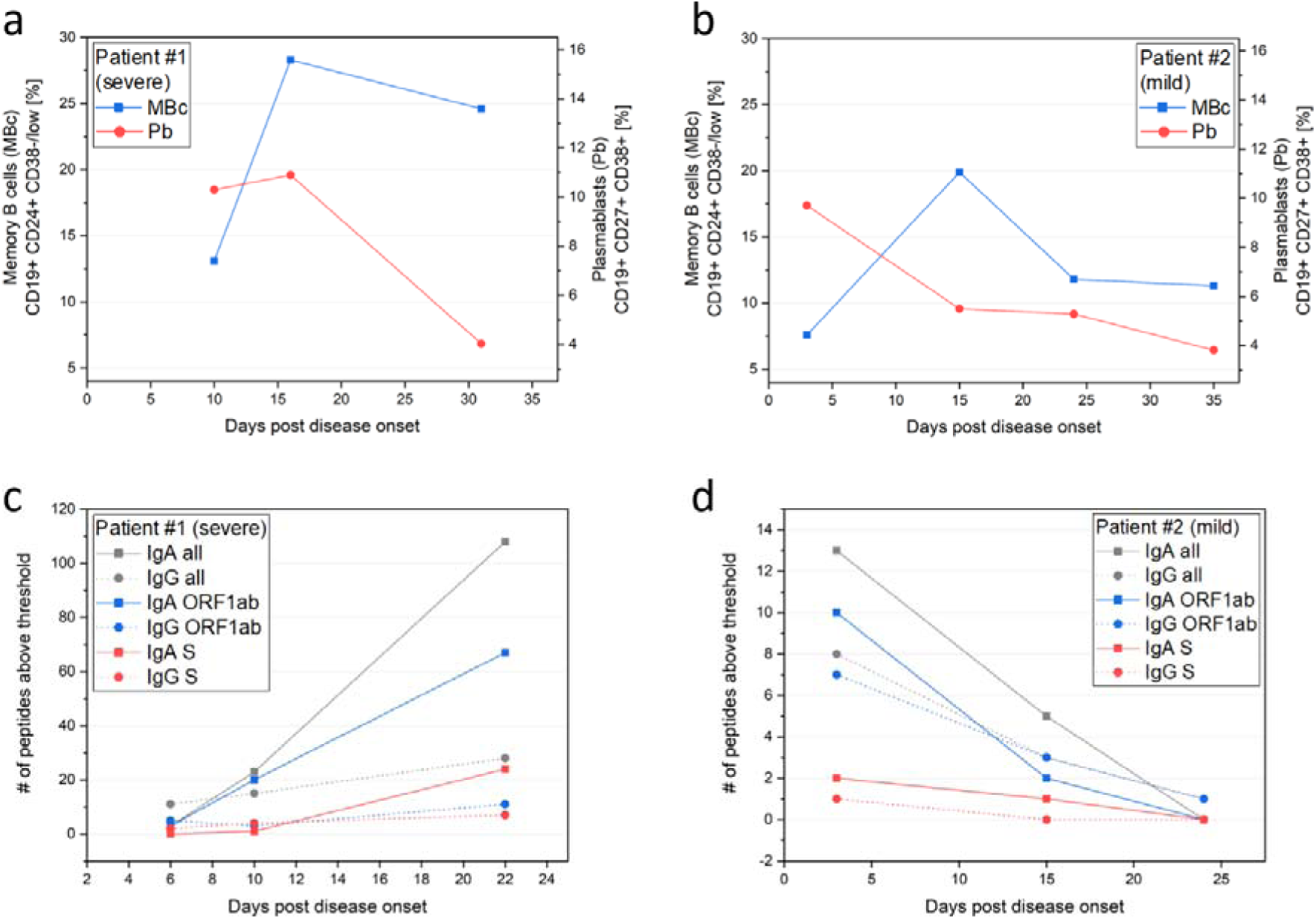
Lymphocyte dynamics and immune response diversity during COVID-19 disease progression. (**a, b**) Patients #1 (more severe case) and #2 (mild case); memory B cells in peripheral blood were stained with CD19, CD24, and CD38, plasmablasts were analyzed by staining CD19, CD27, and CD38. (**c, d**) Overall number of reactive SARS-CoV- 2 peptides in patients #1 (**c**) and #2 (**d**) targeted by IgA and IgG antibodies, identified using full-proteome peptide arrays (see Supporting Information Table S2).

To evaluate the kinetics of B-cell epitopes during the mild and severe course of COVID-19, we used full-proteome peptide microarrays (see Supporting Information for complete microarray data). As a general trend, we observed virus protein-specific IgA and IgG responses with few defined signals, while IgM showed more signals, but without a clear trend. Therefore, we focused on IgA and IgG responses. To define a normal antibody background (negative) on the microarray, we used a healthy donor sample (#5) as a negative control. From sample #5, we selected the 99.9^th^ percentile fluorescence intensity (IgA: 347.8 arbitrary fluorescence units; IgG: 1081.4 arbitrary fluorescence units) as a threshold for positive IgA- and IgG-reactive peptides.

The evolution of IgA and IgG antibodies, targeting SARS-CoV-2 peptides in patients #1 (Fig. 1c) and #2 (Fig. 1d) at different time points shows different dynamics: The more severe case (patient #1) showed few IgA- and IgG-reactive peptides (above control sample threshold) at d6, which considerably increased towards d22 after virus clearance. While reactive IgA peptides at d6 are only found in the ORF1ab polyprotein, the response becomes much broader during disease progression. In comparison, the mild case (patient #2) had a higher number of IgA-reactive SARS-CoV-2 peptides already at d3 post onset of symptoms and showed a decreasing number of reactive peptides from d3 to d24. At this early time point, we already detected defined IgA epitopes in the spike protein, while patient #1 developed IgA targeting spike epitopes only at d22. The trend of early SARS-CoV-2 IgA and IgG antibody response was also observed in the control patient #4 (moderate case, d4 and d12, Supporting Information Figure S1 & S2).

Patients with mild symptoms displayed a much stronger IgA response, soon after onset of symptoms (patient #2 d3, patient #4 d4) that decreased during the course of disease 7 – 14 days later. The IgG response has a similar, but much less pronounced trend and appears more persistent. The number of reactive IgG peptides appears lower in the early acute phase in comparison to IgA.

We analyzed the data in more detail towards individual proteins over time (Fig. 2a–d). On day 6 in patient #1, the IgA antibodies (Fig. 2a) only target the ORF1ab polyprotein. At d10, the IgA response is still low and at d22 it turns into a broad response targeting the spike (S), membrane (M), ORF8, and nucleocapsid (N) proteins. While most IgA ORF1ab signals increase over time in patient #1, three signals decrease considerably. These might have resulted from a non-protective cross-reaction, which are silenced by the immune system. In contrast, in the same patient #1 (Fig. 2c), some IgG responses were already present on d6, targeting the S and M protein. In patient #2 with mild symptoms (Fig. 2b), we observed a stronger and more focused IgA response at d3 against the S, E, N, and ORF1ab proteins compared to patient #1, whereas in the IgG response (Fig. 2d), we only observed one stronger response towards the S protein.

**Figure 2.**
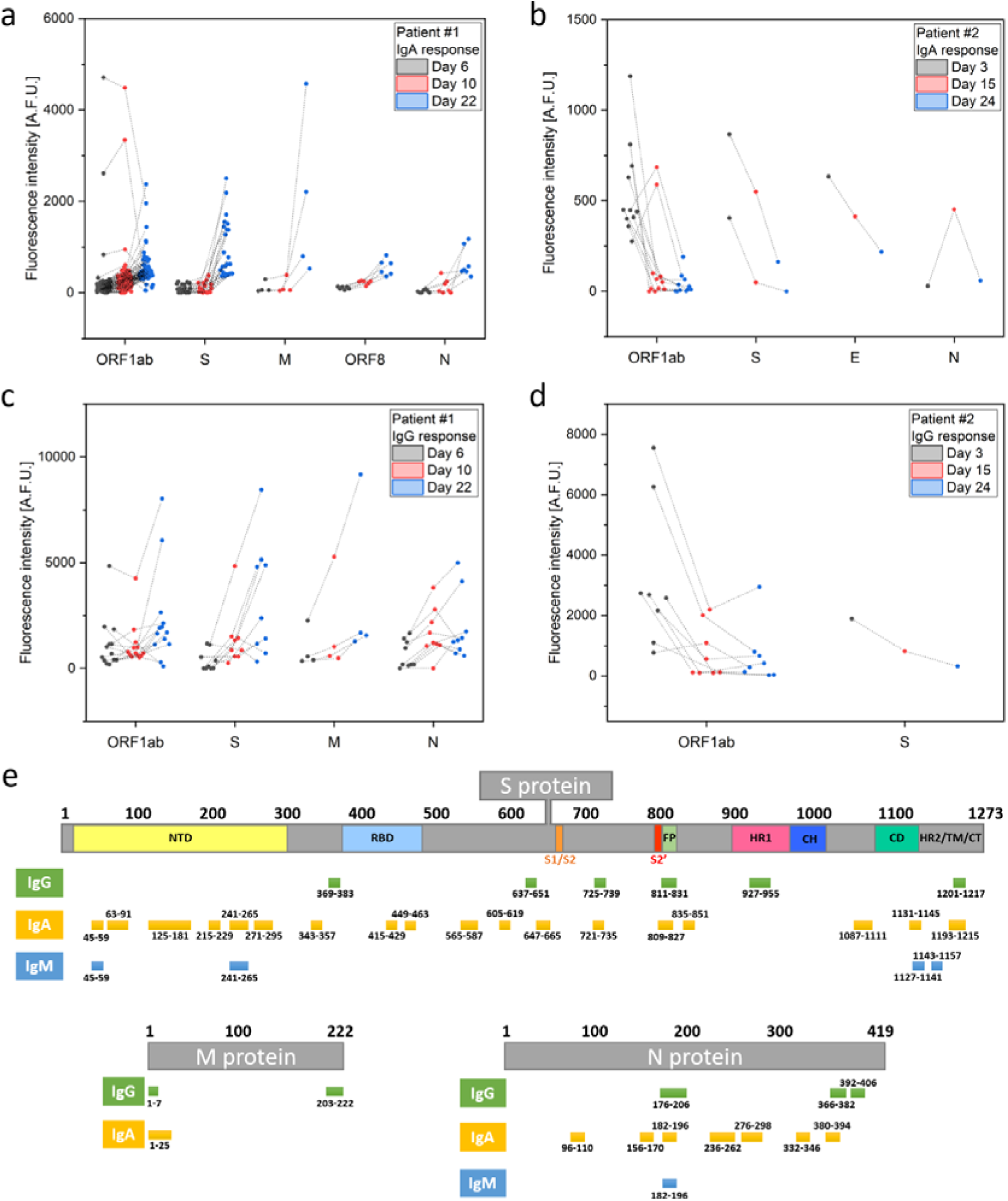
Reactive SARS-CoV-2 peptides following infection. (**a – d**) Evolution of positive responses against SARS-CoV-2 peptides at different time points after onset of disease in patients #1 (more severe symptoms) and #2 (mild symptoms). Data were generated by peptide microarray analyses, IgA (**a, b**) and IgG (**c, d**) signals are shown if at least one time point was above the 99.9^th^ percentile threshold of IgA/IgG signals in healthy control sample #5. Positive signals for ORF1ab polyprotein (ORF1ab), spike glycoprotein (S), envelope protein (E), membrane glycoprotein (M), ORF8 protein, and nucleocapsid phosphoprotein (N) are shown. (**e**) Mapping of reactive IgG, IgA, and IgM signals derived from all nine SARS-CoV-2 patient samples on the S (domains from ^16^), M, and N proteins (see Supporting Information Table S3).

Finally, we visualized the signals/epitopes, derived from all nine patient samples (*vs*. control samples #5) on the S, M, and N proteins (Fig. 2e and Supporting Information Table S3). The data shows generally more IgA than IgG or IgM epitopes, with most epitopes located in the S and N protein.

## Discussion

SARS-CoV-2 has been rapidly spreading among the human population since the end of 2019. Our knowledge of human immune responses to it is limited and the factors that influence symptoms and severity are largely unknown. To address this, we studied the kinetics of B-cell epitope development in relation to the clinical and immunological features of two patients; one experienced a very mild disease course and the other, a more severe. Both patients showed a similar viral load of SARS-CoV-2 following infection (Pfefferle *et al*., submitted). While the proportion of plasmablasts was increased early after symptom onset and then decreased in patient #2 (very mild disease course), the relative plasmablast frequency further increased up to d16 in patient #1 (more severe disease course). After the disease had subsided (d32), the percentage of plasmablasts decreased. Similar observations have been made in a study of a COVID-19 positive female patient, showing a peak of CD19+CD27+CD38+ cells in the blood at day 8 post disease onset, before symptoms were resolved.^8^

Peptide microarrays were employed to obtain a more comprehensive view of B-cell dynamics and a deeper understanding of the development of specific B-cell epitopes during the course of the infection. We investigated the dynamics of IgA, IgG, and IgM antibody responses in very mild compared to more severe SARS-CoV-2 infections during the acute and convalescent phases. The induction of early, strong antibody response was recently reported.^9^ In this study, seroconversion occurred at day 7 post onset of symptoms in 50% of infected individuals. Another study underlined the early responses of IgA, IgM and IgG following SARS-CoV-2 infection.^10^ The authors report a median duration of IgM and IgA antibody detection of 5 days (IQR 3-6). Furthermore, Okba et al.^11^ analyzed IgA and IgG responses in two mild and one severe case using an in-house S1-ELISA. They observed a similar trend with an increase in the IgA response over time in a more severe case. An early IgA response as seen in our patients #2 and #4 was not observed, which might be due to differences in the patients (sample collection dates) or assay performance. Key differences are the limitation of only using S1-proteins for the ELISA (*vs*. whole proteome on the microarray) and a higher sensitivity of peptide arrays.

We detected an early and defined IgA response in patient #2 who had a very mild course of disease. In addition, our patient #4 (mild/moderate course of disease, no hospitalization, see Supporting Information Figures S1 & S2) also showed an early IgA response. IgA is considered a major effector molecule involved in defense mechanisms against viruses and infections with respiratory viruses can induce efficient IgA responses in secretions as well as in sera. Our data suggest that IgA antibodies are valuable diagnostic markers that show strong signals early after onset of mild COVID-19-associated symptoms and that IgA antibodies may be crucial for efficient clearance of SARS-CoV-2.

We identified many spike protein epitopes that are bound by IgA antibodies, which might be crucial for neutralizing viral particles. Early and efficient binding of the spike protein by antibodies can limit viral spread following infection. We also identified several IgA epitopes in the spike protein, such as in the receptor binding domain (AA437–465) and in the fusion peptide domain (AA816–831). In addition, we also confirm parts of a SARS-CoV-2 and SARS-CoV cross-reactive IgG epitope (AA369–383) in the receptor binding domain of spike.^12^

In general, microarrays are valuable tools to identify crucial antibodies and antigens. Recently, COVID-19 patient samples have been analyzed using protein arrays,^13^ as well as with whole-proteome peptide microarrays.^14^ We could confirm some of these epitopes and also identified new epitopes. Since we used a higher number of peptides with more sequence overlap (13 instead of 10 amino acids), our data gives a more comprehensive picture. Furthermore, we show that the IgA response may be highly relevant in disease progression and should be further investigated.

We analyzed a low number of patients, but more are becoming available daily. Our arrays contain exclusively linear peptides and cannot identify antibodies that bind conformational or discontinuous epitopes. The 15-mer peptides, with an overlap of only 13 amino acids maybe too short or the overlap is insufficient to identify all significant antibodies. We considered exclusively the initially published Wuhan strain without mutations, but can quickly incorporate these mutations into the assay, since the array production method is rapid and flexible.^15^ With the limitations listed above, our study contributes to the understanding of differences in the course of disease. More patients need to be included in follow-up studies, to evaluate the importance of the early IgA response and specific epitopes in developing vaccines. Future studies will consider not only seroconvalescent samples, but also the early time points.

Collectively, while antibody dynamics and B-cell epitopes were studied previously, we present the first longitudinal sample analysis of individual B-cell epitope development at early disease phases, comparing degree of disease severity. These data can be the starting point for the development of vaccines or therapeutic antibodies.

## Data Availability

The data that support the findings are available online as supporting information or upon reasonable request from the corresponding authors.

## Acknowledgements

We thank all participants for their involvement in and commitment to this study. We also thank Anneke Novak-Funk and Christina Lehrer for providing input and Anneke Novak-Funk and Elaine Hussey for critically reviewing the manuscript. In addition, we thank Ralf Bischoff, Gregor Jainta, and the company PEPperPRINT for their support.

## Author Contributions

C.D. performed the immunological analyses. J.H. performed all microarray related experiments and analyses. R.K., R.S., and the ID-UKE COVID-19 study group performed clinical monitoring and immunological analyses. F.F.L. supervised the microarray experiments and performed the data analysis. C.D., M.M.A., and F.F.L. supervised the project. C.D., J.H., and F.F.L. wrote the manuscript. All authors helped in developing and revising the manuscript.

## ID-UKE-COVID-19 Study Group

Marylyn M. Addo, Etienne Bartels, Thomas T. Brehm, Christine Dahlke, Anahita Fathi, Monika Friedrich, Svenja Hardtke, Till Koch, Ansgar W. Lohse, My L. Ly, Veronika Schlicker, Stefan Schmiedel, L. Marie Weskamm, Julian Schulze zur Wiesch (University Medical Center Hamburg-Eppendorf)

